# Novel photobiomodulation therapy enhances color discrimination of color vision deficiency due to OPN1LW and/or OPN1MW gene mutations

**DOI:** 10.1101/2023.01.02.22284019

**Authors:** Peihong Wang, Yuqi Wang, Liang Jia

## Abstract

**Purpose:** To investigate the correlations of OPN1LW/OPN1MW (LW/MW) genotypes and clinical phenotypes in individuals with protan/deutan congenital color vision deficiency(CVD), and to explore photobiomodulation (PBM) therapy effects for CVD.

**Design:** Single-center consecutive, retrospective, observational study

**Participants:** 43 subjects (41 males and 2 females) of protan/deutan CVD from the senior ophthalmology department of the PLA General Hospital.

**Methods:** Open-label, single-arm, 4-week pilot trial. Analysis of genetic, clinical, and color vision tests was performed cross-sectionally and longitudinally. Registered with the Chinese Clinical Trial Registry website (ChiCTR2200056761).

**Main Outcome Measures:** types of LW/MW variants, correlation of genotype and phenotype, color discrimination improvements of CVD after PBM therapy.

**Results:** Clinically, the LW gene mutation (8 cases) causes protan CVD, the MW gene mutation (17 cases) and no definite gene mutation (10 cases) cause deutan CVD, dual mutations of LW and MW cause protan (5 cases) or deutan (3 cases) CVD. After individualized therapy, the effects of the MW gene mutation and no definite gene mutation groups are better than those of the LW gene mutation and dual mutations groups.

**Conclusions:** For protan/deutan CVD, PBM therapy can enhance color discrimination, and the result of gene detection is helpful to diagnose the clinical phenotype and predict the therapeutic effects of color vision correction.

**Financial Disclosure(s):** The authors have no proprietary or commercial interest in any of the materials discussed in this article.

---

The cones of the retina are responsible for color sensing and perception. The human retina contains three types of cones that express three kinds of phytochrome molecules: red, green, and blue, which are sensitive to LW, MW, and SW visible light, respectively. However, LW and MW sensitive cones account for most majority (about 94%).^1^

Congenital color vision deficiency (CVD) is a familiar visual dysfunction, and two most common types are protan and deutan CVD. About 8% of men and 0.5% of women worldwide suffer from these two types of CVD.^2,3^ The reason is due to various mutations of OPN1LW/MW genes on the human Xq28 locus^1^, for instance, missense, deletion, duplication, and deletion/insertion. Both LW and MW genes have six exons each, and they are aligned one by one on the X-chromosome^4,5^; on the nucleotide level, they share 96% identity^6^. Normally, the first and sixth exons do not alter^7^. The exon 5 encodes amino acid distinguish that functionally differ Long-wave from Medium-wave sensitive phytochrome^8–10^.

CVD can be categorized by both type and severity. What is the correlation between the mutations of LW/MW genes and the type/severity of CVD? Which type of CVD mutation is more likely to be alleviated after photobiomodulation (PBM) therapy? The two issues raised above are addressed in the following study.

## Methods

This study was approved by the Ethics Committee of PLA General Hospital (Ethical Batch Number: KY2021-017), and was registered on the Chinese Clinical Trial Registry website (Registration Number: ChiCTR2200056761). All participants signed informed consent forms. All experimental protocols were performed per the guidelines of the committee approving the experiments. This study was conducted following the guidelines of the Declaration of Helsinki.

### Participants and Grouping

Participants with protan/deutan CVD were ascertained at Chinese PLA General hospitals from July 2022 to November 2022. Corresponding ophthalmic clinical examinations, color vision tests, and gene sequencing were performed. According to the gene sequencing results, all subjects were divided into four groups: LW gene mutation group, MW gene mutation group, LW/MW genes dual mutations group, and no definite gene mutation group. Protan/deutan CVD was diagnosed clinically with the Farnsworth-Munsell 100 Hue color vision test (FM-100) total error score (TES).

Inclusion criteria: 1) Ishihara’s color blindness test (Ishihara’s) and the FM-100 are used to diagnose congenital protan/deutan CVD in subjects by trained ophthalmologists; 2) Ishihara’s plates are not fully recognized and TES of FM-100 is more than 50; 3) participants’ BCVA was 20/25 or better; 4) participated after agreeing to and signing the informed consent. Exclusion criteria: 1) Patients with other eye diseases that affect vision and color vision, such as glaucoma, cataracts, optic neuritis, and acquired color vision abnormalities; 2) Patients with mental illness and mental abnormalities; 3) Patients with other serious acute diseases (acute myocardial infarction, advanced cancer, etc.).

### Mutation Screening

Blood samples were collected and genomic DNA was isolated from peripheral blood lymphocytes by standard procedures (Omega Bio-Tek Inc, Norcross, GA). Based on targeted exome capture technology, a specific Hereditary Retinal Disease Panel V-5 (personalized customization by MyGenostics Inc, Beijing, China) was used to collect 533 retinal disease-associated genes, and high throughput sequencing was performed with the Illumina HiSeq 2000 platform. The identified variant was confirmed with Sanger sequencing. Segregation analysis was performed when mutations were detected.

### Color Vision Tests

The computer-based FM-100 test detection program^11^ was implemented in a fluorescent lamp room under normative operating procedure recommended by the producer, at a distance of 40 cm and a visual angle of 55°on a high dynamic range (HDR) display measuring 69.8 cm× 36.8 cm with an area of 2569 cm^2^. The program scored the color chess arrangements matrix and drew them on a chart which indicates protan/deutan/tritan color vision dysfunction (Supplementary appendix 1-G), and severity is classified as mild (51<TES<105), moderate (105<TES<191), or severe (TES>191). TES is the parameter for the quantitative evaluation of CVD; the higher the TES, the more serious the CVD.^12^ (Supplementary appendix 1-H)

For quantitative color vision testing, the 38-plate Ishihara’s pseudoisochromatic test (2011 edition, Handaya, Tokyo, Japan) was used. Twenty numeric symbol plates of 2^nd^∼17^th^ and 22^nd^∼25^th^ were applied (Supplementary appendix 1-A∼F). Subjects were asked to identify all numbers on the color blindness test palates in Ishihara’s color blindness book. Each correct answer scored one point, leading to a total score ranging from 0 to 20.

### PBM Therapy

All subjects wore virtual reality glasses (Storm Mirror Little D, Beijing Fengfeng Magic Mirror Technology Co., Ltd.) and got individualized PBM therapy. Subjects with LW gene mutation were exposed to red light irradiation (wavelength 621 nm, full filed illumination, energies density at the cornea was approximately 8.69×10^−4^ mW/cm^2^, duration=6’20’’). Subjects with MW gene mutation and no definite gene mutation were exposed to green light irradiation (wavelength 524nm, full filed illumination, energies density at the cornea was approximately 2.21×10^−3^mW/cm^2^, duration=6’20’’). Subjects with LW/MW dual mutations were exposed to red and green light alternately (the irradiation parameters were the same as in the previous groups, duration=6’20’’). Irradiation frequency was twice daily (morning and evening, with an inter-block period of 12 hours), and lasted four weeks. During irradiation therapy, all participants stare at light and blink normally. All participants were asked to repeat the FM-100 and Ishihara’s tests at the ends of weeks 1, 2, and 4 of therapy. The changes in TES and Ishihara’s score were statistically analyzed.

### Statistical Analysis

The primary outcome was the FM-100 TES, and the secondary outcomes were Ishihara’s color blindness test score. Shapiro-Wilk tests were used to check the distribution of data and the homogeneity of variance was checked using Levene’s test. Continuous variables were described using mean & standard deviation (normally distributed data) and median & inter-quartile range (non-parametric data). The therapy effect was analyzed using the Kruskal-Wallis test, followed by the LSD-t test. The effect of weeks of therapy was analyzed using the repeated measurement ANOVA followed by Wilcoxon matched-pairs signed-rank test. Bonferroni post hoc adjustments were used for multiple comparisons. Analyses were performed using R 4.2.1 and P < 0.05 was considered statistically significant.

## Results

### Demographic Data, Genetic Analysis, and Color Vision Test Results

43 subjects (41 males and 2 females; age range 7∼55 years; mean age 27.3 years) participated in this study. Subjects were divided into four groups according to the type of gene mutation (Figure 1): LW gene mutation group (8 males, 18.6%), MW gene mutation group (17 males, 39.53%), LW/MW dual mutations group (7 males, 1 female, 18.6%) and no definite gene mutation group (9 males, 1 female, 23.26%). Information on sex, age, clinical diagnosis, TES, and Ishihara’s score of subjects in 4 groups was detailed in table 1.

**Figure 1:**
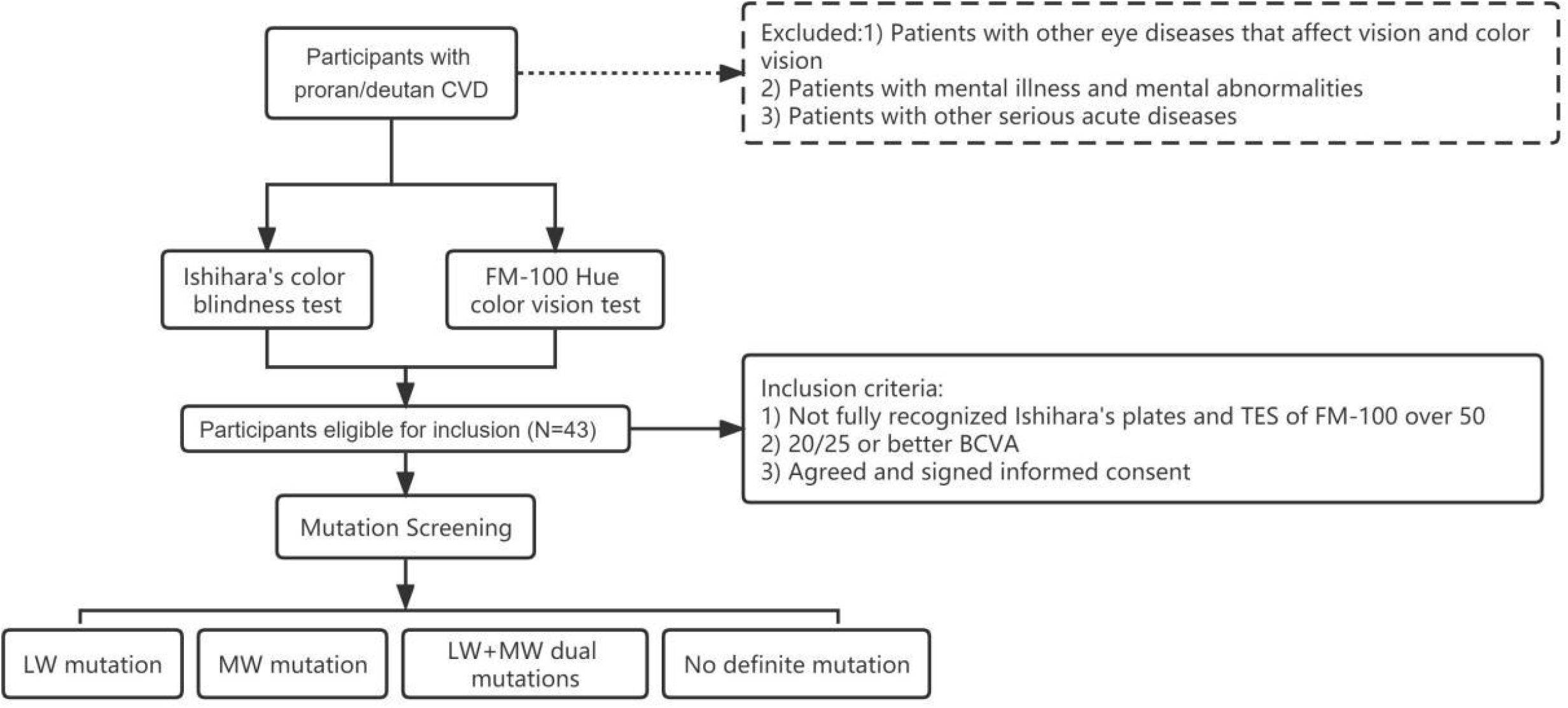
Consort diagram of PBM therapy study for protan/deutan CVD.

**Table 1.**
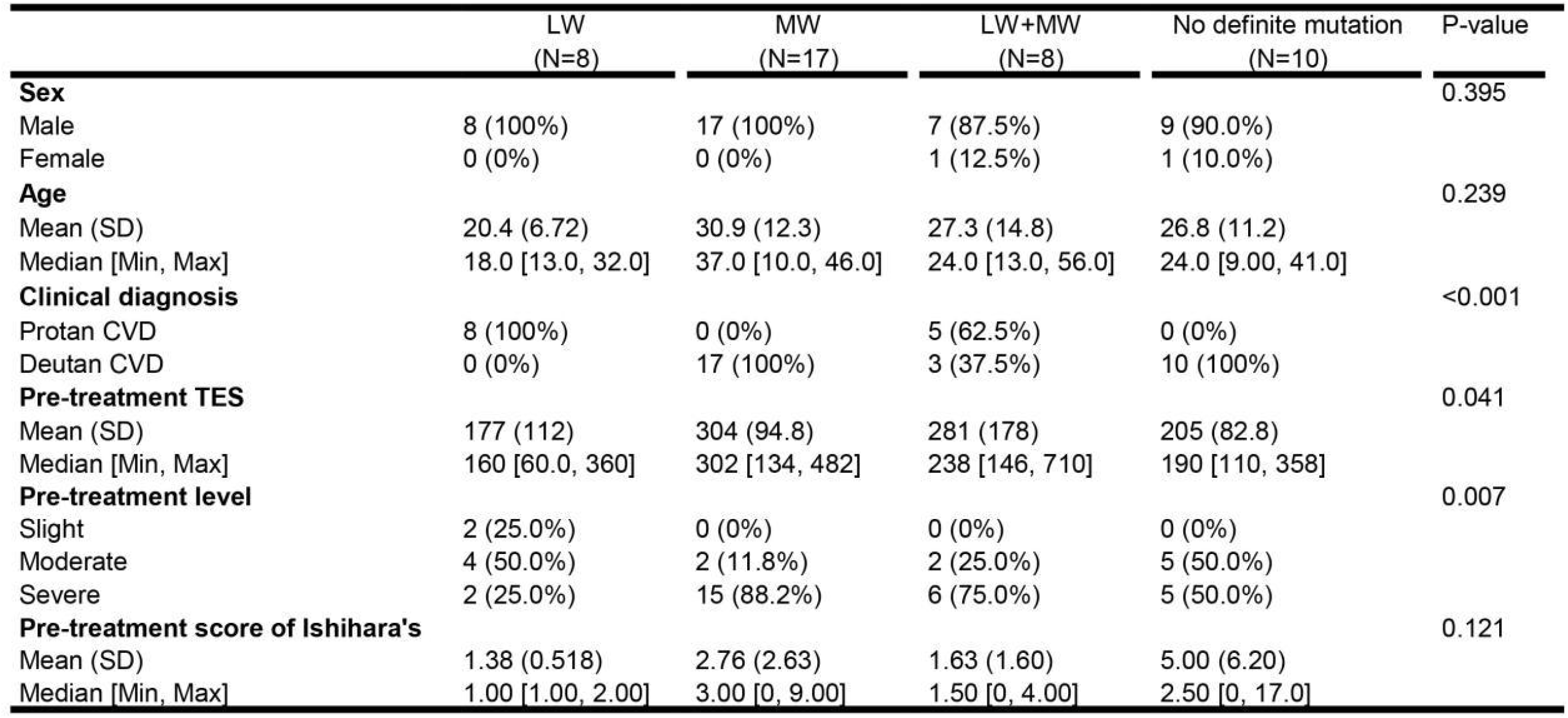
General information and color vision test results of four groups

### Correlations of LW/MW Genotypes and Clinical Phenotypes

Clinically, subjects with LW gene mutation are protan CVD; subjects with MW gene mutation and no definite gene mutation are deutan CVD; subjects with LW+MW dual mutations are protan or deutan CVD. (Detailed in Table 1) The proportions of different types of exons hemizygous deletion (HD) in the first three groups are shown in Figure 2-A∼D.

**Figure 2.**
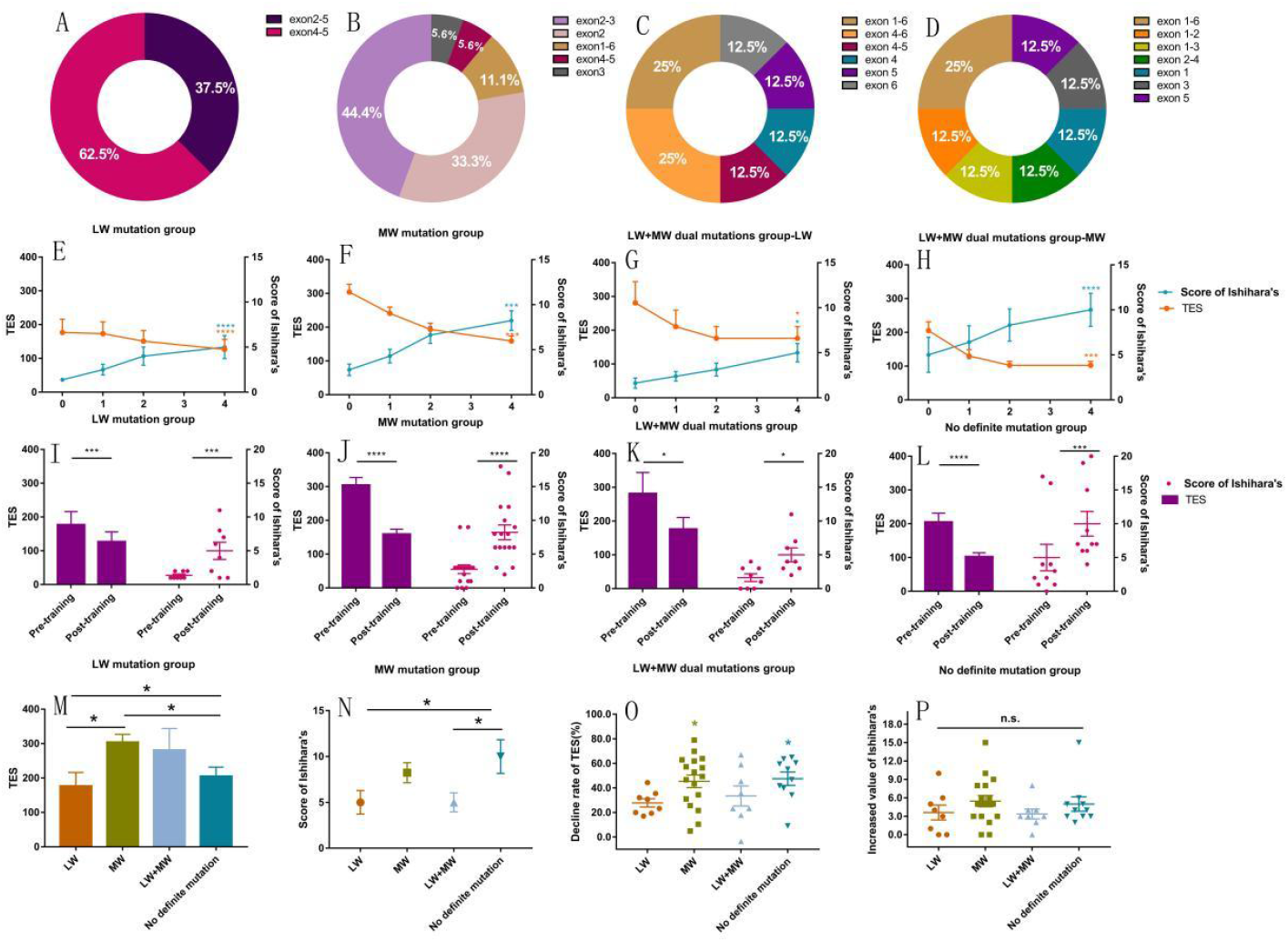
LW/MW genotypes and PBM therapy effects of four groups: LW mutation, MW mutation, LW+MW dual mutations, and no definite gene mutation groups. A∼D The proportions of different types of exons hemizygous deletion (HD) in the first three groups; E∼H In four groups, TES decreased gradually with time, and the difference was significant compared with that pre-therapy; Ishihara’s score increased gradually with time, and the difference was significant compared with that pre-therapy; I∼L, O, and P MW gene mutation group had the best effect, and the LW+MW dual mutations group had the worst effect; M MW gene mutation group had the highest TES, which was significantly different from LW gene mutation and no definite gene mutation groups pre-therapy; N No definite gene mutation group had the highest Ishihara’s score, which was significantly different from the LW gene mutation and LW+MW dual mutation groups. * *P*<0.05, ** *P*<0.01, *** *P*<0.005, \*\*\*\**P*<0.001.

### Comparison of TES and Ishihara’s Score of Four Groups’ Pre-therapy

By comparing the TES of four groups’ pre-therapy, it could be found that the MW gene mutation group had the highest score, which was significantly different from the LW gene mutation and no definite gene mutation groups. (Figure 2-M) By comparing Ishihara’s score of four groups’ pre-therapy, it could be found that no definite gene mutation group had the highest score, which was significantly different from the LW gene mutation and LW+MW dual mutation groups. (Figure 2-N)

### Changes in TES and Ishihara’s Score During the 4-week Therapy

In four groups, TES decreased gradually with time, Ishihara’s score increased gradually with time, and significant differences were observed(Figure 2-E∼H). According to the TES and Ishihara’s score changes of four groups pre-therapy and post-therapy, it could be found that the MW gene mutation group had the best effect, and the LW+MW dual mutations group had the worst effect. (Figure 2-I∼L, 2-O, P).

## Discussion

In mammals, only primates have complex trichromatic color vision^13^, for example, humans can perceive red, green, and blue (three primary colors). In addition, trichromatic vision (routine trichromacy) is normal in both males and females of humans and non-human primates14. The physiological basis of human trichromatic visions is cones with three kinds of phytochrome: L, M, and S cones^15^. The LW gene encodes the phytochrome responsible for red perception, while the MW gene encodes the phytochrome responsible for green perception^16^. Some combinations of individual single nuclear acid polymorphisms (SNPs) are related to photoreceptor functional disorder of cones, and exchange modification is viewed as a significant cause of color vision loss.^1,17–19^

Various clinical color vision examinations for CVD include quantitative and qualitative methods, such as the Nagel anomaloscope and FM-100^20^, and so on. Among them, the Nagel anomaloscope is generally considered the “gold standard” for color vision tests^21^, however, its tedious application has prevented it from gaining widespread popularity. FM-100 is the most commonly used method in clinics. Ghose^22^ used the digital FM-100 as one of the methods of color vision inspection and compared the results with the conventional FM-100. The results demonstrated that the digital FM-100 reduced the test time by more than 50%. Digital FM-100 is also superior to conventional FM-100 in terms of sensitivity and specificity. This research confirms that the digital FM-100 has the advantages of simple, fast operation and strong professional function of test results. In this study, we also used a computer-based FM-100 test detection program on an HDR display to make sure the color chess of FM-100 could be presented truly (details in the methods section).

Ishihara’s color blindness test is a classic color vision screening tool, and it lacks the function of quantifying color perception. Considering that these color blindness plates were designed for various types of subjects with CVD, we made an ingenious modification to enable it to quantify the ability of color perception indirectly and roughly. We selected all 20 number symbol plates from the test book (details in the methods section). One point is awarded if the subject correctly identifies all of the numbers on the plate, and 20 plates correspond to 20 points. In this way, each person’s color vision discrimination ability is quantified as a score range of 0∼20. This is a useful change that is recommended for clinicians.

In this study, combining the mutation type of exon with clinical examination, it was found that all subjects with LW gene mutation were protan CVD clinically. All subjects with MW gene mutation and those without mutation detected were deutan CVD clinically. Part of the subjects with LW/MW dual mutations were protan CVD clinically, and the rest were deutan CVD clinically. The genotype can not be extrapolated from the phenotype, for example, the genotype of protan CVD may be LW mutation or LW/MW dual mutations. On the contrary, the genotype also may not be representative of the phenotype, for example, LW/MW dual mutations may lead to protan or deutan CVD. However, when the two methods are combined to address the precision diagnosis and therapy, dominance and high certainty are revealed. Interestingly, subjects without definite mutation still suffer abnormal color vision and are all clinically diagnosed with deutan CVD. When the gene mutation in the Xq28 region is in this arrangement: LW-LW-MW, that is, the locus of the MW gene is inserted by a complete LW gene, the subsequent MW gene will not be translated, leading to the expression defect of green phytochrome^1^. Due to the limitations of detection methods and conditions, when the subject’s gene sequence is in the above conditions, and no mutation such as exon deletion occurs, the specific arrangement of the subject cannot be determined. The resolution of this issue requires further study.

In previous studies, CVD was considered a stable birth defect. In addition to genetic therapy^23,24^, which is still in the preclinical phase, there are currently no effective therapeutic measures available for this population. In this study, the mutation type of exon was combined with the clinical therapy effects, and it was found that: first, the TES of FM-100 decreased and scores of Ishihara’s increased significantly after therapy, which indicated that the subject’s color discrimination may be improved; second, we used PBM therapy, which is a non-invasive physical method, and the possible principles are illustrated with a schematic diagram in Figure 3. Unlike gene therapy, which is currently being studied, it poses no risk and can be carried out and tracked anytime and anywhere. Third, the therapy effect of subjects with different types of gene mutations varies, and it is gratifying that the therapy effect of subjects with deutan CVD (high incidence) was better than that of subjects with protan CVD (low incidence).

**Figure 3.**
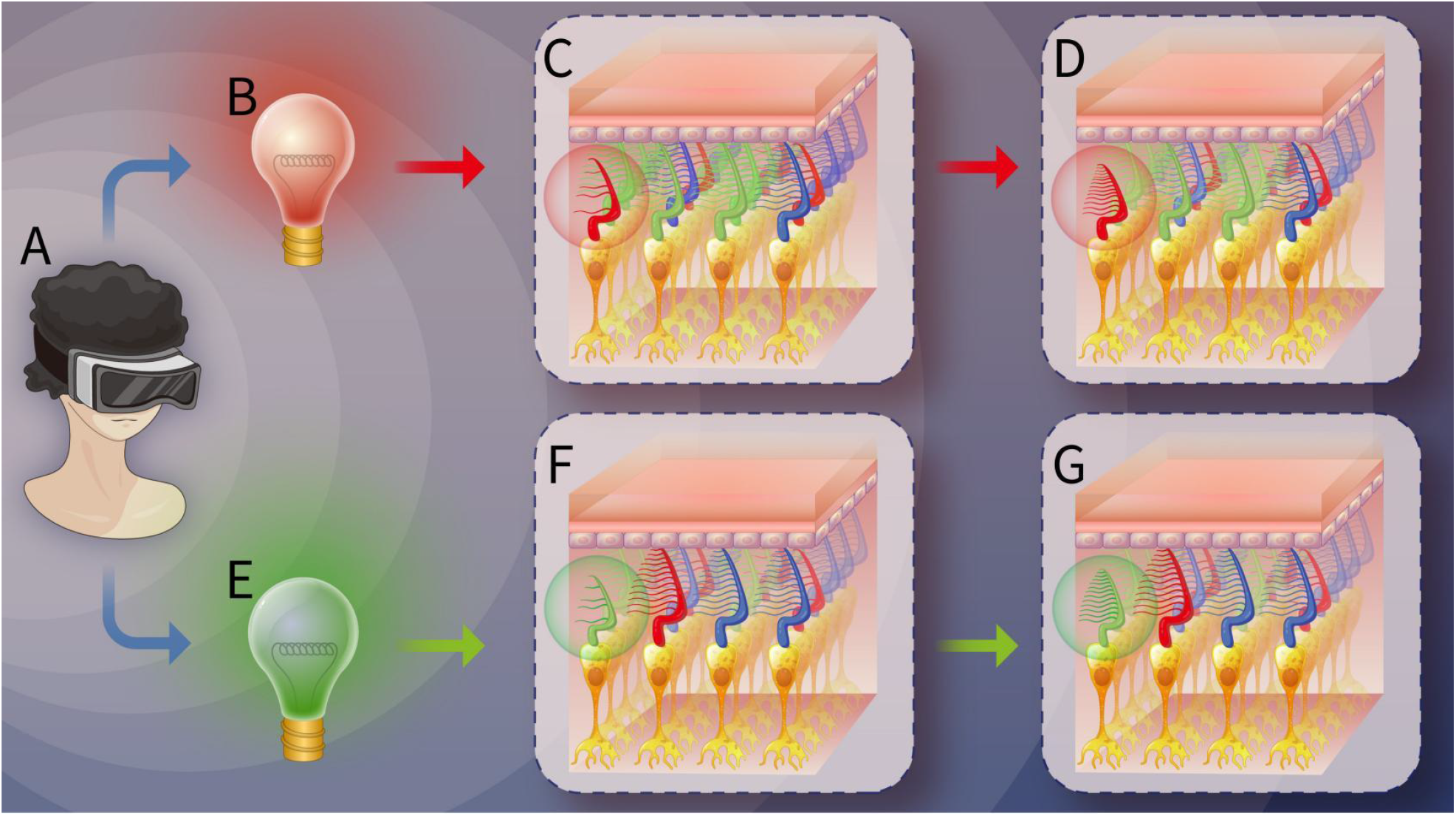
Schematic diagram of the possible principle of the improvement of color vision discrimination ability for patients with protan/deutan CVD during photobiomodulation(PBM) therapy. A: Patient wearing VR head display for therapy; B Light source of red light PBM therapy; C The outer segment of red cones are congenitally deficient in patients with protan CVD; D After red light therapy, the morphology of red cones was improved, the content of red phytochrome protein in the outer segment was increased, and the red perception ability was improved, but were still not reached the morphology and ability of normal green and blue cones; E Light source of green light PBM therapy; F The outer segment of green cones are congenitally deficient in patients with deutan CVD; G After green light therapy, the morphology of green cones was improved, the content of green phytochrome protein in the outer segment was increased, and the green perception ability was improved, but still not reached the morphology and ability of normal red and blue cones.

To summarize, gene sequencing is worth popularizing as an accurate method for identifying specific gene mutation types. Through the results of gene detection, we can not only speculate the clinical type and severity of CVD individuals but also predict the effect and outcome of color vision correction therapy, which provides strong clinical support for the diagnosis and therapy of CVD.

## Data Availability

All data produced in the present study are available upon reasonable request to the authors

## Acknowledgment

The authors would like to thank Zhibing Li, and Shengyong Dong (From Chinese PLA General Hospital) for their support of the clinical trial, Guanyu Che, Zhonghua Geng, Yuanxia Zhang, Juan Wang, and Qing Deng for their assistance in recruiting volunteers.

## Author contributions

Peihong Wang: Study Design, Data Collection, Data Analysis, Writing - Original draft. Yuqi Wang: Data Analysis, Figures Composition. Liang Jia: Study Design, Data Collection, Writing - Review, and editing.

## Abbreviations/Acronyms

CVD: Color Vision Deficiency
LW: OPN1LW
MW: OPN1MW
PBM: Photobiomodulation
FM-100: Farnsworth-Munsell 100 Hue Color Vision Test
Ishihara’s: Ishihara’s color blindness test HDR = High Dynamic Range
VR: Virtual Reality

## Reference

1. Neitz M, Neitz J. Intermixing the OPN1LW and OPN1MW Genes Disrupts the Exonic Splicing Code Causing an Array of Vision Disorders. Genes 2021;12:1180–1198. Available at: https://www.mdpi.com/2073-4425/12/8/1180 [Accessed November 2, 2022].

2. Tian Y, Tang H, Kang T, et al. Inverse-Designed Aid Lenses for Precise Correction of Color Vision Deficiency. Nano Letters 2022;22.

3. Tsekouras GE, Rigos A, Chatzistamatis S, et al. A novel approach to image recoloring for color vision deficiency. Sensors 2021;21.

4. J N, D T, Ds H. Molecular genetics of human color vision: the genes encoding blue, green, and red pigments. Science (New York, NY) 1986;232:193–202. Available at: https://pubmed.ncbi.nlm.nih.gov/2937147/ [Accessed November 2, 2022].

5. D V, J N, Rw D. Tandem array of human visual pigment genes at Xq28. Science (New York, NY) 1988;240:1669–1672. Available at: https://pubmed.ncbi.nlm.nih.gov/2837827/ [Accessed November 2, 2022].

6. J N, Tp P, Rl E, et al. Molecular genetics of inherited variation in human color vision. Science (New York, NY) 1986;232:203–210. Available at: https://pubmed.ncbi.nlm.nih.gov/3485310/ [Accessed November 2, 2022].

7. Winderickx J, Battlsti L, Hlbiya Y, et al. Haplotype diversity in the human red and green opsin genes: Evidence for frequent sequence exchange in exon 3. Human Molecular Genetics 1993;2.

8. Neitz M, Neitz J, Jacobs GH. Spectral tuning of pigments underlying red-green color vision. Science 1991;252:971–974.

9. Davidoff C, Neitz M, Neitz J. Genetic testing as a new standard for clinical diagnosis of color vision deficiencies. Translational Vision Science and Technology 2016;5:2–18.

10. Neitz M, Neitz J. Variety of photopigment genes underlying red-green colour vision. In: ; 1997:33–43.

11. Farnsworth, Munsell. Farnsworth-Munsell 100 Hue color vision test. Available at: https://www.colour-blindness.com/farnsworth-munsell-100-hue-colour-vision-test/.

12. J B. A practical guide for colour-vision examination: report of the Standardization Committee of the International Research Group on Colour-Vision Deficiencies. Ophthalmic & physiological optics : the journal of the British College of Ophthalmic Opticians (Optometrists) 1985;5:265–285. Available at: https://pubmed.ncbi.nlm.nih.gov/3876538/ [Accessed November 2, 2022].

13. Neitz M, Patterson SS, Neitz J. Photopigment genes, cones, and color update: disrupting the splicing code causes a diverse array of vision disorders. Curr Opin Behav Sci 2019;30:60–66. Available at: https://www.ncbi.nlm.nih.gov/pmc/articles/PMC7081934/ [Accessed November 4, 2022].

14. Jacobs GH. Evolution of colour vision in mammals - PMC. Available at: https://www.ncbi.nlm.nih.gov/pmc/articles/PMC2781854/ [Accessed November 4, 2022].

15. Sahu DK, Panda SP, Meher PK, et al. Construction, De-Novo Assembly and Analysis of Transcriptome for Identification of Reproduction-Related Genes and Pathways from Rohu, Labeo rohita (Hamilton). PLoS One 2015;10:e0132450. Available at: https://www.ncbi.nlm.nih.gov/pmc/articles/PMC4509579/ [Accessed November 4, 2022].

16. Thoreson WB, Dacey DM. Diverse Cell Types, Circuits, and Mechanisms for Color Vision in the Vertebrate Retina. Physiol Rev 2019;99:1527–1573. Available at: https://www.ncbi.nlm.nih.gov/pmc/articles/PMC6689740/ [Accessed November 4, 2022].

17. Neitz J, Neitz M. The genetics of normal and defective color vision - PubMed. Available at: https://lib.plagh.cn/s/gov/nih/nlm/ncbi/pubmed/G.https/21167193/ [Accessed November 4, 2022].

18. Anon. De novo intrachromosomal gene conversion from OPN1MW to OPN1LW in the male germline results in Blue Cone Monochromacy - PubMed. Available at: https://lib.plagh.cn/s/gov/nih/nlm/ncbi/pubmed/G.https/27339364/?;x-chain-id=7vwx3atsg8ow [Accessed November 4, 2022].

19. J C, M N, H H, et al. Functional photoreceptor loss revealed with adaptive optics: an alternate cause of color blindness. Proceedings of the National Academy of Sciences of the United States of America 2004;101:8461–8466. Available at: https://pubmed.ncbi.nlm.nih.gov/15148406/ [Accessed November 4, 2022].

20. Azuma K, Inoue T, Fujino R, et al. Comparison between blue-on-yellow and white-on-white perimetry in patients with branch retinal vein occlusion. Sci Rep 2020;10:20009–20017. Available at: https://www.ncbi.nlm.nih.gov/pmc/articles/PMC7672051/ [Accessed November 4, 2022].

21. Taore A, Lobo G, Turnbull PR, Dakin SC. Diagnosis of colour vision deficits using eye movements. Sci Rep 2022;12:7734–7748. Available at: https://www.ncbi.nlm.nih.gov/pmc/articles/PMC9095692/ [Accessed November 4, 2022].

22. Ghose S, Parmar T, Dada T, et al. A new computer-based Farnsworth Munsell 100-hue test for evaluation of color vision. Int Ophthalmol 2014;34:747–751. Available at: http://link.springer.com/10.1007/s10792-013-9865-9 [Accessed November 2, 2022].

23. Neitz M, Neitz J. Curing Color Blindness--Mice and Nonhuman Primates. Cold Spring Harbor Perspectives in Medicine 2014;4:a017418–a017418. Available at: http://perspectivesinmedicine.cshlp.org/lookup/doi/10.1101/cshperspect.a017418 [Accessed November 5, 2022].

24. Mancuso K, Neitz M, Hauswirth WW, et al. Long-Term Results of Gene Therapy for Red-Green Color Blindness in Monkeys. Investigative Ophthalmology & Visual Science 2010;51:1423–1433.

