## supplementary appendix 2 for "Novel photobiomodulation therapy enhances color discrimination of color vision deficiency due to OPN1LW and/or OPN1MW gene mutations"

**Table 1: The examination results before and at the end of week 1, week 2 and week 4 of LW gene mutation group.**

| **Number** | **Sex** | **Age ranges** | **Type of CVD** | **Farnsworth-Munsell 100 Hue color vision test** | | | | | **Ishihara's test score** | | | | |
| --- | --- | --- | --- | --- | --- | --- | --- | --- | --- | --- | --- | --- | --- |
|  |  |  |  | **W0** | **W1** | **W2** | **W4** | **Decline Rate** | **W0** | **W1** | **W2** | **W4** | **Increased Value** |
| 1 | male | 10~19 | Protan | 108 | 132 | 124 | 60 | 44.44% | 2 | 4 | 8 | 7 | 5 |
| 2 | male | 20~29 | Protan | 324 | 298 | 264 | 220 | 32.10% | 1 | 1 | 1 | 1 | 0 |
| 3 | male | 10~19 | Protan | 62 | 80 | 64 | 50 | 19.35% | 2 | 4 | 6 | 8 | 6 |
| 4 | male | 10~19 | Protan | 60 | 78 | 58 | 48 | 20.00% | 2 | 4 | 5 | 6 | 4 |
| 5 | male | 30~39 | Protan | 180 | 156 | 128 | 116 | 35.56% | 1 | 4 | 7 | 11 | 10 |
| 6 | male | 10~19 | Protan | 164 | 158 | 142 | 136 | 17.07% | 1 | 2 | 3 | 4 | 3 |
| 7 | male | 10~19 | Protan | 360 | 348 | 308 | 276 | 23.33% | 1 | 1 | 1 | 2 | 1 |
| 8 | male | 10~19 | Protan | 108 | 132 | 124 | 60 | 44.44% | 2 | 4 | 8 | 7 | 5 |

**Table 2: The examination results before and at the end of week 1, week 2 and week 4 of MW gene mutation group.**

| **Number** | **Sex** | **Age ranges** | **Type of CVD** | **Farnsworth-Munsell 100 Hue color vision test** | | | | | **Ishihara's test score** | | | | |
| --- | --- | --- | --- | --- | --- | --- | --- | --- | --- | --- | --- | --- | --- |
|  |  |  |  | **W0** | **W1** | **W2** | **W4** | **Decline Rate** | **W0** | **W1** | **W2** | **W4** | **Increased Value** |
| 1 | male | 10~19 | Deutan | 482 | 324 | 276 | 244 | 49.38% | 3 | 5 | 7 | 8 | 5 |
| 2 | male | 20~29 | Deutan | 450 | 356 | 320 | 196 | 56.44% | 0 | 0 | 1 | 2 | 2 |
| 3 | male | 10~19 | Deutan | 366 | 256 | 160 | 132 | 63.93% | 1 | 3 | 5 | 6 | 5 |
| 4 | male | 10~19 | Deutan | 206 | 244 | 204 | 196 | 4.85% | 1 | 4 | 5 | 9 | 8 |
| 5 | male | 30~39 | Deutan | 362 | 240 | 120 | 76 | 79.01% | 3 | 2 | 3 | 3 | 0 |
| 6 | male | 10~19 | Deutan | 186 | 142 | 96 | 56 | 69.89% | 9 | 12 | 15 | 17 | 8 |
| 7 | male | 10~19 | Deutan | 302 | 180 | 124 | 112 | 62.91% | 3 | 5 | 5 | 6 | 3 |
| 8 | male | 10~19 | Deutan | 234 | 220 | 187 | 174 | 25.64% | 3 | 3 | 5 | 8 | 5 |
| 9 | male | 10~19 | Deutan | 238 | 240 | 216 | 156 | 34.45% | 9 | 10 | 11 | 12 | 3 |
| 10 | male | 20~29 | Deutan | 206 | 100 | 96 | 88 | 57.28% | 0 | 1 | 4 | 6 | 6 |
| 11 | male | 10~19 | Deutan | 294 | 184 | 138 | 112 | 61.90% | 3 | 5 | 13 | 18 | 15 |
| 12 | male | 10~19 | Deutan | 290 | 248 | 186 | 164 | 43.45% | 3 | 3 |  | 3 | 0 |
| 13 | male | 30~39 | Deutan | 374 | 320 | 286 | 258 | 31.02% | 0 | 4 | 7 | 10 | 10 |
| 14 | male | 10~19 | Deutan | 330 | 264 | 212 | 176 | 46.67% | 1 | 2 | 4 | 6 | 5 |
| 15 | male | 10~19 | Deutan | 342 | 384 | 344 | 268 | 21.64% | 3 | 5 | 6 | 6 | 3 |
| 16 | male | 10~19 | Deutan | 374 | 244 | 200 | 176 | 52.94% | 3 | 7 | 9 | 12 | 9 |
| 17 | male | 10~19 | Deutan | 134 | 156 | 126 | 120 | 10.45% | 2 | 2 | 6 | 8 | 6 |

**Table 3: The examination results before and at the end of week 1, week 2 and week 4 of LW+MW dual mutations group.**

| **Number** | **Sex** | **Age ranges** | **Type of CVD** | **Farnsworth-Munsell 100 Hue color vision test** | | | | | **Ishihara's test score** | | | | |
| --- | --- | --- | --- | --- | --- | --- | --- | --- | --- | --- | --- | --- | --- |
|  |  |  |  | **W0** | **W1** | **W2** | **W4** | **Decline Rate** | **W0** | **W1** | **W2** | **W4** | **Increased Value** |
| 1 | Female | 20~29 | Deutan | 242 | 88 | 80 | 80 | 66.94% | 4 | 4 | 5 | 7 | 3 |
| 2 | male | 50~59 | Deutan | 170 | 192 | 176 | 176 | -3.53% | 1 | 2 | 1 | 4 | 3 |
| 3 | male | 40~49 | Deutan | 146 | 132 | 112 | 112 | 23.29% | 3 | 5 | 7 | 11 | 8 |
| 4 | male | 10~19 | Protan | 258 | 232 | 212 | 212 | 17.83% | 0 | 1 | 2 | 2 | 2 |
| 5 | male | 10~19 | Protan | 230 | 176 | 148 | 148 | 35.65% | 0 | 1 | 2 | 4 | 4 |
| 6 | male | 20~29 | Protan | 234 | 128 | 96 | 96 | 58.97% | 3 | 2 | 2 | 3 | 0 |
| 7 | male | 10~19 | Protan | 710 | 532 | 388 | 388 | 45.35% | 0 | 1 | 2 | 3 | 3 |
| 8 | male | 20~29 | Protan | 258 | 206 | 198 | 198 | 23.26% | 2 | 3 | 4 | 6 | 4 |

**Table 4: The examination results before and at the end of week 1, week 2 and week 4 of no definite gene mutation group.**

| **Number** | **Sex** | **Age ranges** | **Type of CVD** | **Farnsworth-Munsell 100 Hue color vision test** | | | | | **Ishihara's test score** | | | | |
| --- | --- | --- | --- | --- | --- | --- | --- | --- | --- | --- | --- | --- | --- |
|  |  |  |  | **W0** | **W1** | **W2** | **W4** | **Decline Rate** | **W0** | **W1** | **W2** | **W4** | **Increased Value** |
| 1 | Female | 40~49 | Deutan | 194 | 72 | 68 | 68 | 64.95% | 3 | 2 | 4 | 7 | 4 |
| 2 | male | 0~9 | Deutan | 186 | 128 | 112 | 112 | 39.78% | 2 | 3 | 5 | 7 | 5 |
| 3 | male | 10~19 | Deutan | 298 | 220 | 118 | 118 | 60.40% | 4 | 5 | 5 | 6 | 2 |
| 4 | male | 20~29 | Deutan | 110 | 98 | 100 | 100 | 9.09% | 17 | 17 | 19 | 20 | 3 |
| 5 | male | 30~39 | Deutan | 358 | 228 | 160 | 160 | 55.31% | 1 | 1 | 4 | 4 | 3 |
| 6 | male | 10~19 | Deutan | 134 | 96 | 88 | 88 | 34.33% | 0 | 6 | 10 | 15 | 15 |
| 7 | male | 30~39 | Deutan | 278 | 178 | 148 | 148 | 46.76% | 2 | 5 | 7 | 6 | 4 |
| 8 | male | 10~19 | Deutan | 178 | 86 | 72 | 72 | 59.55% | 4 | 5 | 6 | 9 | 5 |
| 9 | male | 30~39 | Deutan | 206 | 128 | 120 | 120 | 41.75% | 1 | 3 | 5 | 7 | 6 |
| 10 | male | 20~29 | Deutan | 110 | 68 | 40 | 40 | 63.64% | 16 | 17 | 18 | 19 | 3 |
