## Supplementary figures and images for "Novel photobiomodulation therapy enhances color discrimination of color vision deficiency due to OPN1LW and/or OPN1MW gene mutations"

### supplementary appendix 1

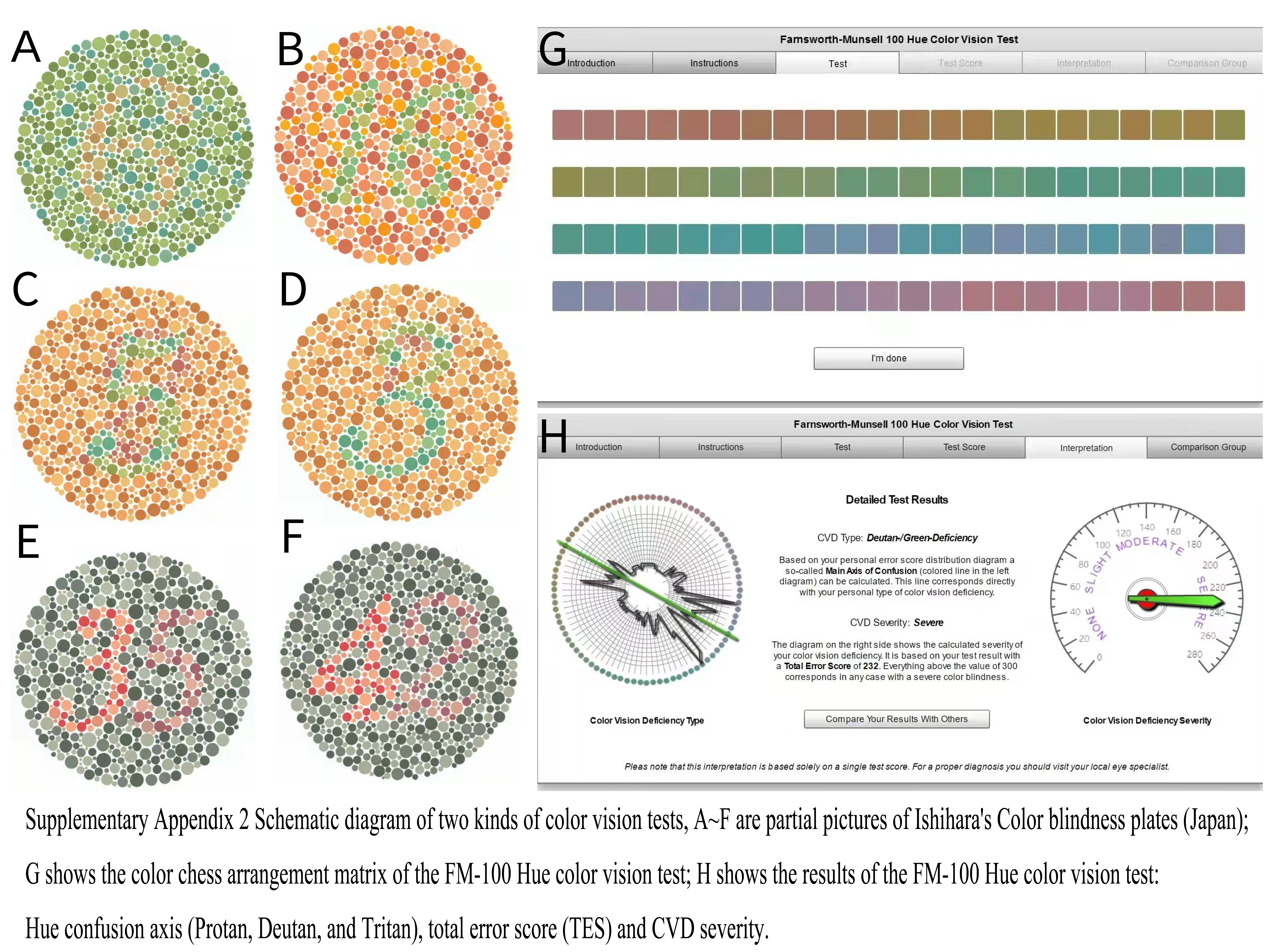
